# Development of a reporting guideline for umbrella reviews on epidemiological associations using cross-sectional, case-control, and cohort studies: the Preferred Reporting Items for Umbrella Reviews of Cross-sectional, Case-control, and Cohort studies (PRIUR-CCC)

**DOI:** 10.1101/2022.12.28.22283572

**Authors:** Marco Solmi, Kelly D Cobey, David Moher, Sanam Ebrahimzadeh, Elena Dragioti, Jae Il Shin, Joaquim Radua, Samuele Cortese, Beverley Shea, Nicola Veronese, Lisa Hartling, Michelle Pollock, Matthias Egger, Stefania Papatheodorou, John P.A. Ioannidis, Andre F. Carvalho

## Abstract

**Introduction:** Observational studies are fraught with several biases including reverse causation and residual confounding, which may limit the credibility of reported associations. Overview of reviews of observational studies (i.e., umbrella reviews) synthesize systematic reviews with or without meta-analyses of cross-sectional, case-control, and cohort studies, and may also aid in the grading of the credibility of reported associations. The number of published umbrella reviews has been increasing at a rapid pace. Recently, a reporting guideline for overviews of reviews of healthcare interventions (PRIOR, Preferred Reporting Items for Overviews of Reviews) was published, but the field lacks reporting guidelines for umbrella reviews of observational studies. Thus, our aim is to develop a reporting guideline for umbrella reviews on cross-sectional, case-control, and cohort studies assessing epidemiological associations.

**Methods and Analyses:** We will adhere to established guidance on how to develop reporting guidelines in health research and follow four steps to prepare a PRIOR extension for systematic reviews of cross-sectional, case-control, and cohort studies testing epidemiological associations between an exposure and an outcome, namely Preferred Reporting Items for Umbrella Reviews of Cross-sectional, Case-control, and Cohort studies (PRIUR-CCC).

Step 1 will be the project launch to identify stakeholders. Step 2 will be a literature review of available guidance to conduct umbrella reviews. Step 3 will be a Delphi study sampling authors and editors of umbrella reviews, Delphi surveys and checklists of epidemiological studies, as well as funders, practitioners, and policy makers, which will be conducted in three rounds. Step 4 will encompass the finalization of PRIUR-CCC statement, including a checklist, a flow diagram, explanation, and elaboration document. Deliverables of each step will be as follows. First, identifying stakeholders to involve according to relevant expertise and end-user groups, with an equity, diversity, and inclusion lens. Second, completing a narrative review of methodological guidance on how to conduct umbrella reviews, a narrative review of methodology and reporting in published umbrella reviews, and preparing an initial PRIUR-CCC checklist for Delphi study Round 1. Third, preparing a PRIUR-CCC checklist with guidance after Delphi study. Fourth, publishing and disseminating PRIUR-CCC statement.

**Ethics and Dissemination:** PRIUR-CCC will guide reporting of umbrella reviews on epidemiological associations, with the aim to improve quantitative, credible, and transparent reporting, in a field of evidence synthesis where there is important methodological heterogeneity of reviews, and where sources of bias in original observational studies can lead to misleading conclusions.

**Strengths:**

1. This is the first protocol for reporting guidance of umbrella reviews of epidemiological associations
2. This protocol follows the guidance for reporting checklist, which are standard in the field.
3. This protocol is urgently needed given the large number of umbrella reviews on epidemiological associations emerging across different branches of science

## Introduction

There is evidence that the number of systematic reviews and meta-analyses in the literature has increased geometrically over the past two decades^1,2^. Due to the increasing number of systematic reviews and meta-analyses on a given topic over the years, the field of knowledge synthesis has developed systematic reviews of systematic reviews, also called “reviews of reviews”, “overviews of (systematic) reviews”, “meta-reviews”, or “umbrella reviews” ^3–8^. Umbrella reviews can include interventional studies or observational studies^9^. Overviews of reviews and umbrella reviews ideally aim to provide a comprehensive and systematic synthesis following the steps of a systematic review (i.e., literature search, methodological quality appraisal, quantitative analysis where feasible and appropriate, etc.) with systematic reviews as the unit of analysis ^2,9,10^. The field has seen a sharp increase in the number of published overviews of reviews and umbrella reviews over the past decade. For example, just limiting to umbrella reviews, approximately 56 were published in 2010, whilst 560 (10 times increase in yearly publications) were published in 2021 (PubMed, [(umbrella review]).

There is important variability between and within the approach of overviews of reviews and umbrella reviews, making results hardly comparable^11–13^. This heterogeneity is not surprising, considering the large heterogeneity in the conception and implementation of both systematic reviews and meta-analyses^14,15^. The methodology used to conduct systematic reviews and meta-analyses, as well as the quality of reporting, can affect conclusions and potentially lead to misleading interpretation of findings, misinforming policy makers, professional organizations and regulatory bodies, practitioners, patients, the public, and other stakeholders. The quality and credibility of evidence synthesis efforts is also largely based on the quality of the credibility of their unit of inclusion (i.e., individual studies). There is consensus that randomized controlled trials start from a higher credibility in the evidence-based medicine pyramid, and lower credibility is assigned to observational studies, which are more prone to bias. Interventional studies are not free from limitations, but in general experimental designs such as randomized controlled trials can protect from a number of biases, such as confounding by indication, or reverse causality. By contrast, observational evidence is prone to these and other biases, including excess of significance bias^16^. Among observational studies, different study designs are adapted depending on the research question to be answered. For instance, studies measuring prognostic accuracy or prediction models are typically cohort studies that need internal development of the model, internal and external validation, calibration and accuracy measures^17^. Cross-sectional studies can instead be used to investigate biomarkers or diagnostic accuracy of a given construct/test, or the prevalence of a disease. Other research questions, and typically epidemiological associations between two factors are generally explored with cohort, case-control, and cross-sectional studies^18,19^. More specifically, among studies investigating epidemiological associations, cross-sectional studies are typically used to measure the association between two factors, neglecting the direction of such association, while case-control and cohort studies are frequently used to measure associations between a construct of interest, and putative risk factors^1,14,20–23^, or its outcomes^24,25^, with the exposure occurring before the outcome. Research questions for which umbrella reviews of observational studies can be used are reported in Table 1.

**Table 1.** Frequent research questions for which overviews of reviews of observational studies are used.

| **Research questions** | **Appropriate study design** |
| --- | --- |
| **Epidemiologic associations covered by PRIUR-CCC** |  |
| What are the risk/protective factors for a disease?<br>(e.g. what are factors that increase or decrease the risk of developing multiple sclerosis?) | Case-control, cohort studies |
| What are the outcomes associated with a risk factor?<br>(e.g. what are the outcomes of being exposed to a traumatic event?) | Case-control, cohort studies |
| What is the association between two factors/entities?<br>(e.g. is schizophrenia associated with more frequent substance use?) | Cross-sectional |
| **Other observational research questions and designs** |  |
| What is the prevalence or incidence of a disorder? | Cross-sectional, cohort studies |
| What is the diagnostic accuracy of different tests/thresholds for the same condition, or across multiple disorders? | Cross-sectional studies |
| What are the prognostic or diagnostic multivariate prediction models for a given disorder, or multiple disorders? | Cohort studies |

Reporting guidelines, defined as “a checklist, flow diagram, or explicit text to guide authors in reporting a specific type of research, developed using explicit methodology”^26^, can be useful to improve the transparency, quality, and reporting of individual studies, reviews, or umbrella reviews. Interventional and observational evidence pose different methodological quality and reporting challenges, that are reflected from different reporting guidance for the two study designs. For observational evidence several checklists of essential information to be reported in a paper, are available for individual studies, systematic reviews, and meta-analyses. For observational studies, the Enhancing the QUAlity and Transparency Of health Research (EQUATOR) Network has disseminated Standards for Reporting Diagnostic accuracy studies (STARD 2015)^27^, Transparent reporting of a multivariable prediction model for individual prognosis or diagnosis (TRIPOD)^28^, and Strengthening the Reporting of Observational Studies in Epidemiology (STROBE)^29^. For systematic reviews, the Preferred Reporting Items of Systematic Reviews and Meta-Analyses 2020 (PRISMA 2020)^30^ is a broad guide inclusive of a range of primary study designs, and more specific statements are available, i.e. the PRISMA for Diagnostic Test Accuracy (PRISMA-DTA)^31^, and PRISMA for reviews including harms outcomes (PRISMA harms)^32^, or the guidance on conducting systematic reviews and meta-analyses of observational studies on etiology (COSMOS-E)^33^ (Table 2). The different checklists addressing different study designs and research questions well reflect their different methodological challenges, from original research to evidence synthesis.

**Table 2.** Key reporting guidelines across different research questions that can be addressed with observational studies.

| Research questions | Primary studies | Systematic Reviews |
| --- | --- | --- |
| **Epidemiologic associations covered by PRIUR-CCC** |  |  |
| Epidemiological associations | STROBE<sup>29</sup>, RECORD<sup>60</sup> | MOOSE<sup>61,62</sup>, PRISMA 2020<sup>30</sup>,<br>PRISMA harms<sup>32</sup>, COSMOS-E<sup>33</sup> |
| **Other observational research questions and designs** |  |  |
| Diagnostic accuracy study | STARD 2015<sup>27</sup> | PRISMA-DTA<sup>31</sup> |
| Multivariate prediction models for individual prognosis or diagnosis | TRIPOD<sup>28</sup> | PRISMA 2020<sup>30</sup> |

*Legend. COSMOS-E, guidance on conducting systematic reviews and meta-analyses of observational studies on etiology; EQUATOR, Enhancing the QUALity and Transparency Of health Research ; MOOSE<sup>61,62</sup>, Meta-analysis Of Observational Studies in Epidemiology; PRIUR, Preferred Reporting Items for Umbrella Reviews; PRIUR-CCC, PRIUR for cross-sectional, case-control, cohort studies; PRISMA<sup>30,32</sup>, Preferred Reporting Items of Systematic Reviews and Meta-Analyses; PRISMA-DTA<sup>31</sup>, PRISMA for Diagnostic Test Accuracy; RECORD<sup>60</sup>, REporting of studies Conducted using Observational Routinely-collected Data; RoB, risk of bias; STARD<sup>27</sup>, Standards for Reporting Diagnostic accuracy studies; STROBE<sup>29</sup>, Strengthening the Reporting of Observational Studies in Epidemiology; TRIPOD<sup>28</sup>, Transparent reporting of a multivariable prediction model for individual prognosis or diagnosis.*

Regarding overview of reviews and umbrella reviews, virtually all reporting checklists proposed so far have focused on interventional evidence. The first proposed checklist^34^ was based on AMSTAR^35^ and Cochrane guidance^36^. The same year, one further checklist for overviews of reviews^37^ merged evidence from PRISMA for abstracts^38^, the PRIO checklist^39^, and the Overview Quality Assessment Questionnaire^40^. Then, a checklist for overviews of reviews ^41^ was developed based on Cochrane recommendations^36^, and the older versions of PRISMA^42^ (and not PRISMA 2020^30^), and A MeaSurement Tool to Assess systematic Reviews) quality assessment tool (AMSTAR)^35,43^ (and not AMSTAR 2^44^). Later, checklist was developed for systematic reviews of reviews including harms, called Preferred reporting items for overviews of systematic reviews (PRIO)^39^, based on an older version of PRISMA^42^, PRISMA harms^32^, and PRISMA for systematic review protocols^45^. The same group also published a checklist for abstract of overviews of reviews of healthcare interventions^46^. Recently the Preferred Reporting Items for Systematic Reviews of Systematic Reviews/Meta-analyses (PRIOR) statement has been published to guide reporting of overviews of reviews of health interventions^3^, adhering to EQUATOR guidance^26^. In PRIOR protocol^3^, authors acknowledged that several relevant sources exist that provide guidance on overviews of reviews or umbrella reviews, but they did not adhere to guidance endorsed by the EQUATOR Network.

Regarding umbrella reviews (i.e., observational evidence investigating epidemiological associations), to the best of our knowledge no EQUATOR-adherent guidance has been developed, registered with, or disseminated by EQUATOR group, nor any specific checklist has been previously proposed. Given that PRIOR focuses on interventional evidence, and that different reporting guidelines are needed for observational evidence (cross-sectional, case-control, cohort studies) on epidemiological associations versus interventional evidence, the aim of this project is to develop evidence-based and agreement-based guideline PRIOR-extension for reporting umbrella reviews (i.e. cross-sectional, case-control, cohort studies testing epidemiological associations), adhering to established guidance^26^, and building on PRIOR statement^3^. Specifically, this project will yield a PRIOR extension for umbrella reviews of cross-sectional, case-control, and cohort studies (PRIUR-CCC), which will be published in a peer-reviewed journal and available via a dedicated website.

## Methods

### Transparency statement

We have also submitted this protocol to The Ottawa Health Science Network-Research Ethics Board and have obtained consent (20220639-01H). All participants to the Delphi survey will give informed consent, which they will be able to withdraw at any time (yet anonymous responses can’t be withdrawn). This protocol is publicly available at medrxiv.org (MEDRXIV/2022/283572).

All study data and materials, will be publicly available.

### Study Design

This study will follow EQUATOR guidance for developing reporting checklists^26^, and will be composed on four steps, namely project launch, literature review, Delphi survey, and guideline statement preparation (Figure 1).

**Figure 1.**
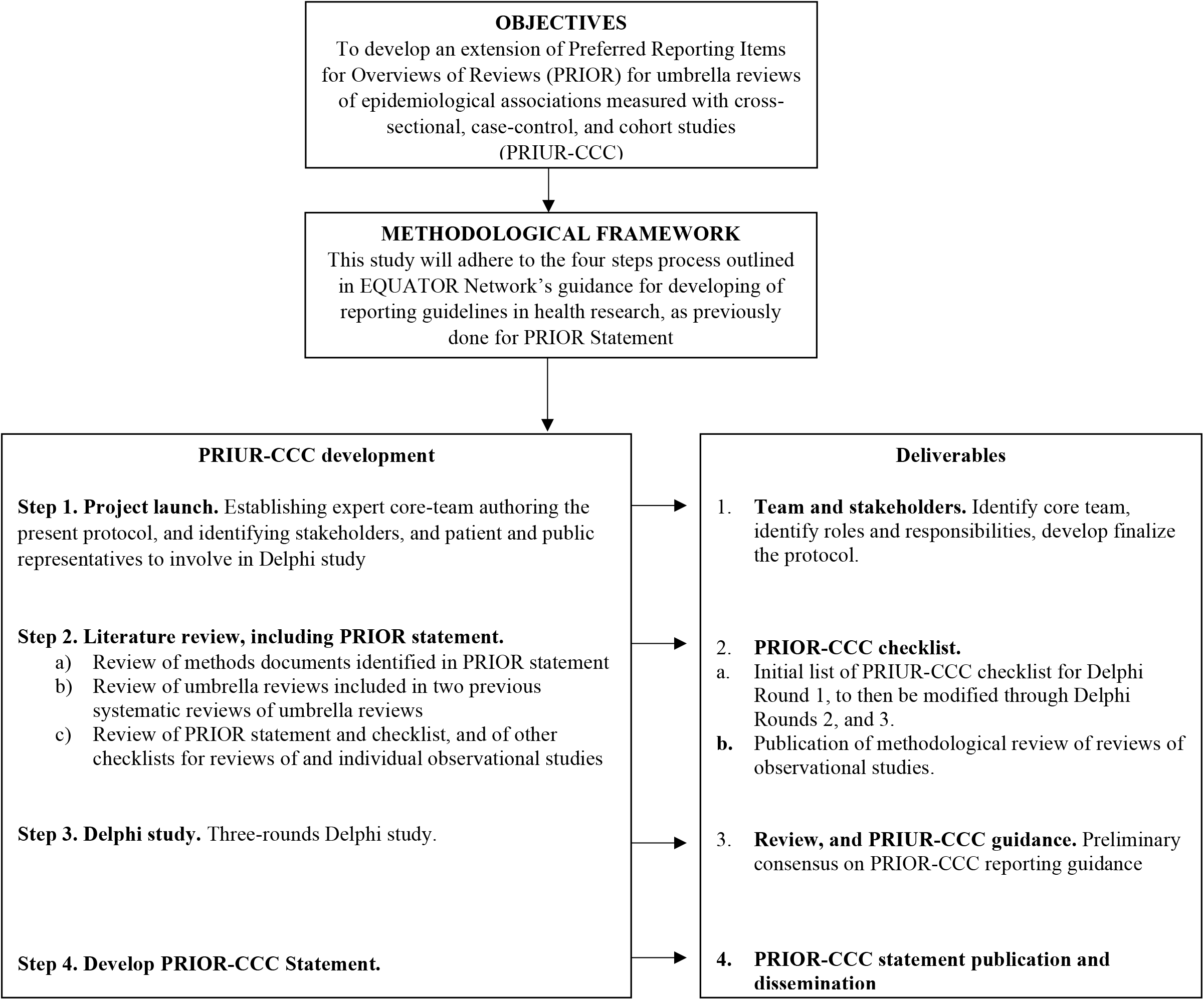
Development of PRIUR-CCC statement flow diagram. *Legend. EQUATOR, Enhancing the QUAlity and Transparency Of health Research* ; *PRIOR*, Preferred Reporting Items for Overviews of Reviews; PRIUR-CCC, PRIOR for cross-sectional, case-control, cohort studies

#### Project launch

Project launch will consist of reaching an agreement on roles and responsibilities of core team members (e.g., identifying stakeholders that will participate in the Delphi study). A core group of researchers have prepared the protocol of the project. These authors will be the core team of the project, and have extensive record of umbrella reviews on cross-sectional, case-control, and cohort studies investigating epidemiological associations. The project’s day-to-day steps and its finalization will be responsibility of shared first authors, and last author. All other authors will contribute to the protocol, literature review, initial set of checklist items, Delphi survey, and final PRIUR-CCC statement. The identification and involvement of stakeholders will follow the Practical Guidance for Involving Stakeholders in Health Research from the Multi Stakeholder Engagement (MuSE) Consortium^47^. Reporting of involvement of patients and public will adhere to Guidance for Reporting Involvement of Patients and the Public 2 (GRIPP2)^48^.

#### Literature narrative review

We will review the systematic reviews conducted in the context of PRIOR development regarding available guidance to conduct umbrella reviews. Then, since two systematic reviews of umbrella reviews have already been conducted by the members of this project ^2,12^, those two systematic reviews will be used to assess methodology and reporting of included umbrella reviews. Given that PRIUR-CCC will not focus on diagnostic test accuracy and prediction models, we will not cover those study designs as they are out of scope for PRIUR-CCC. After having reviewed PRIOR documentation and having reviewed the two previous systematic reviews of umbrella reviews, we will extract key methodological factors from identified umbrella reviews, and publish a review of methodology and reporting of previous umbrella reviews. The findings of this review will be used to identify PRIOR domains and items where changes are needed, and to produce an initial PRIOR checklist to start the Delphi study with.

#### Delphi survey

We will conduct a Delphi study. Delphi studies use social science survey techniques to structure communication between participants in order to drive consensus and make a collective decision^49^. Typically, Delphi studies use several rounds of surveys in which participants, vote on issues. Between rounds, results of voting are aggregated and anonymized. They are then presented back to participants along with their own individual scores, and feedback on why others voted as they did^50,51^. Among others, two strengths of this method include allowing for effective communication, limiting direct confrontation between individuals, and giving participants the opportunity to consider the group’s thoughts and to compare and adjust their own score in the next round. ***Participants***. We will aim to include 100 participants in Delphi study round 1, as done in PRIOR development^3^. Participants to the Delphi study will meet any of the following criteria; a) they have publication track of umbrella reviews in various fields, or b) have publication track in Delphi surveys, or c) have publication track of reporting checklists of cross-sectional, case-control, and cohort studies, or editors of peer-reviewed journals that published umbrella reviews on observational evidence or that have interest in umbrella reviews, or e) funders of research or meta-research, or f) practitioners, g) policy makers, h) evidence synthesis associations (e.g. Cochrane, Campbell collaboration, Joanna Briggs Institute, others)^3^. Participants will be recruited via a two-step process. First, we have established a core group of participants based on criteria a) or b) or c) described above (project launch), and past solid collaboration track record, that are authoring the present protocol. Second, additional stakeholders will be invited, according to a) to h) criteria above. ***Delphi study methods***. This Delphi study will consist of three rounds. For the first two Delphi rounds, we will ask participants to complete an online survey which will be administered using Google Forms, structuring forms based on purpose-built platform for Delphi survey development and management^52^. The third round will consist of a facilitated online consensus group meeting (using Zoom™^53^).

##### Round 1

Age, gender, geographical area (https://www.who.int/countries) and stakeholders group will be collected anonymously. Similarly to what has been done for scoping reviews^54^, we will build on existing PRIOR statement, which parallels PRISMA 2020^30^ statement, for consistency and continuity with existing established reporting guidance for evidence syntheses. All participants will be asked what items of PRIOR^3^ will have to i) remain unchanged, ii) what will have to be changed and how, iii) what will have to be removed (three answer options, one possible choice). In addition, participants will have the possibility to propose new items. As a starting point, the core team composed of authors of this protocol will provide a set of suggested items for participants to vote on (Appendix 1). This starting set of items will be built based on experience in umbrella reviews. Participants will respond indicating their agreement/disagreement to have each item included in the PRISMA-CCC reporting guideline. Participants will be provided with a free text box to fill in with additional comments to explain why they voted how they did or to propose wording amendments to the item, or new items. To decide to keep or remove a PRIOR item, or to add a new item, will require a minimum of 80% consensus among participants (based on findings from a systematic review of Delphi studies^55^). Items where an 80% consensus has not been reached, as well as changes to PRIOR items and new items will be voted on again in Round 2. We anticipate the survey will take about 20 minutes to complete and will provide participants with a 3-week window to take part, with reminders sent after 1 and 2 weeks, respectively. We will pilot test the survey among the authors of the present protocol.

##### Round 2

All participants who completed round 1 of the Delphi will be re-invited to take part in round 2. Items which achieved consensus in round 1 will be shared with the participants. We will then ask participants to re-vote on any items that did not reach consensus or that were newly suggested. When re-voting on items that appeared in round 1, participants will also be provided with all comments provided by participants to justify their responses. Again, an 80% of consensus will be used to determine what items to include/exclude from the PRIUR-CCC guideline. If a new proposed item will be overlapping with existing items, Delphi moderator will add a comment to the new proposed item pointing to existing overlapping item.

##### Round 3

The core group and a purposeful selection of participants will be invited to round 3. We will aim to ensure representation from each of our geographically diverse institutions, each of the stakeholder groups, and demographic variables including gender. Based on previous Delphi surveys and feasibility, we will invite no more than 30 participants in total to ensure feasibility to Round 3^56,57^. Round 3 will be moderated by core group members authoring the protocol, who will rotate every two items. We will present all participants with the results (i.e., frequency of responses for each item, comments, changes, and new items) of round 2 of the Delphi prior to the meeting and summarize these again briefly at the start of the consensus meeting. Participants will have the opportunity to discuss outstanding items one-by-one. They will then be asked to vote anonymously using real-time voting technology available via Zoom on each of these items. While diverse time zones will present challenges in other ways a virtual meeting may foster equity, diversity, and inclusion of participants who might otherwise not have had funding or capacity to travel to an in-person event.

##### Analysis

For completers and drop-outs, demographics, and responses will be presented using descriptive statistics using SPSS28. We will identify which items have and have not reached consensus for inclusion or exclusion based on our definition of 80% agreement and report this information for each Delphi round. The list of items identified for inclusion in PRIUR-CCC will be collated after round 3 and we will report the outcomes of participants ranking of these items in a table.

#### Timeline

The project has not started yet, and will be started in January 2023 and completed by December 2023.

#### Guidance statement

Co-first and last authors of this protocol will prepare the first draft of final PRIUR-CCC statement, that will then be approved after re-iteration with other authors. This PRIUR-CCC statement will include the report of the whole project, the PRIUR-CCC checklist, the PRIUR-CCC flow diagram, and an explanatory document that will inform on how to use PRIUR-CCC, with examples. PRIUR-CCC statement will be published in peer-reviewed journals, and on dedicated platforms. The Delphi process reporting will be informed by the Conducting and REporting of DElphi Studies (CREDES) checklist^58^ (Table 3).

**Table 3.** CREDES checklist for survey studies.

| **Items of reporting** | **Reported on page** |
| --- | --- |
| *Purpose and rationale.* The purpose of the study should be clearly defined and demonstrate the appropriateness of the use of the Delphi technique as a method to achieve the research aim. A rationale for the choice of the Delphi technique as the most suitable method needs to be provided. |  |
| *Expert panel.* Criteria for the selection of experts and transparent information on recruitment of the expert panel, sociodemographic details including information on expertise regarding the topic in question, (non)response and response rates over the ongoing iterations should be reported. |  |
| *Description of the methods.* The methods employed need to be comprehensible; this includes information on preparatory steps (How was available evidence on the topic in question synthesised?), piloting of material and survey instruments, design of the survey instrument(s), the number and design of survey rounds, methods of data analysis, processing and synthesis of experts' responses to inform the subsequent survey round and methodological decisions taken by the research team throughout the process. |  |
| *Procedure.* Flow chart to illustrate the stages of the Delphi process, including a preparatory phase, the actual 'Delphi rounds', interim steps of data processing and analysis, and concluding steps. |  |
| *Definition and attainment of consensus.* It needs to be comprehensible to the reader how consensus was achieved throughout the process, including strategies to deal with non-consensus. |  |
| *Results.* Reporting of results for each round separately is highly advisable in order to make the evolving of consensus over the rounds transparent. This includes figures showing the average group response, changes between rounds, as well as any modifications of the survey instrument such as deletion, addition or modification of survey items based on previous rounds. |  |
| *Discussion of limitations.* Reporting should include a critical reflection of potential limitations and their impact of the resulting guidance. |  |
| *Adequacy of conclusions.* The conclusions should adequately reflect the outcomes of the Delphi study with a view to the scope and applicability of the resulting practice guidance. |  |
| *Publication and dissemination.* PRIUR-CCC will be published in peer-reviewed journals, with details on methodology (including CREDES checklist) in main text or supplementary material. |  |

*Legend. CREDES, Conducting and REporting of DELphi Studies; PRIOR, Preferred Reporting Items for Overviews of Reviews; PRIUR-CCC, PRIOR for cross-sectional, case-control, cohort studies*

### Patient and public involvement

Reporting of involvement of patients and public will adhere to Guidance for Reporting Involvement of Patients and the Public 2 (GRIPP2)^48^. All authors completing all three rounds of Delphi process and the core group will be invited to review and finally co-author the publication.

## Discussion

PRIUR-CCC will provide a reporting framework to guide future umbrella reviews of observational studies assessing epidemiological associations, that can be used from researchers, reviewers, funders, and editors to evaluate the transparency and quality of reporting of umbrella reviews, across different research questions. We propose that having reporting guidelines is of crucial relevance when included studies follow an observational design, with high baseline risk of confounding factors and numerous sources of bias potentially guiding misleading results^11,59^. We aim to publish the report of this project including the final PROR-CCC guidelines.

## Data Availability

All study data and materials, including the protocol, will be publicly available on the Open Science Framework (https://osf.io/cpty5) as well as in the published paper(s).

## Author Contributions

MS, KC, LH, and AFC drafter the first version of the protocol. All authors revised multiple times the protocol and approved the final version.

## Funding Statement

LH is supported by a Canada Research Chair in Knowledge Synthesis and Translation.

This research received no other specific grant from any funding agency in the public, commercial or not-for-profit sectors

## Competing Interest Statement

MS received honoraria/has been a consultant for Angelini, Lundbeck, Otsuka. SCo declares honoraria and reimbursement for travel and accommodation expenses for lectures from the following non-profit associations: Association for Child and Adolescent Central Health (ACAMH), Canadian ADHD Alliance Resource (CADDRA), and the British Association of Pharmacology (BAP) for educational activity on ADHD.

All other co-authors have no conflict of interest to declare.

## Notes

### Clinical Protocols

https://osf.io/cpty5

### Author Declarations

We have submitted this protocol to The Ottawa Health Science Network- Research Ethics Board and have obtained consent (20220639-01H).

